# Summer of Translational Aging Research for Undergraduates (STAR U): Short-term outcomes of a training program to advance diversity in aging research

**DOI:** 10.1101/2024.05.23.24307742

**Authors:** Kiana K. Chan, Jennifer J. Manly, Stephanie A. Cosentino, Adam M. Brickman

## Abstract

**Purpose:** The Summer of Translational Aging Research for Undergraduates (STAR U) program, funded by the National Institute on Aging and the Alzheimer’s Association, aims to advance diversity in aging research through undergraduate education. Here, we evaluate the effectiveness of the program in cultivating a diverse cohort of scientists from underrepresented backgrounds.

**Method:** Forty-eight (96%) of 50 STAR U alumni completed a program evaluation survey between April and August 2023. The survey collected data on demographic characteristics of the alumni, educational/career goals, program experiences, and post-program outcomes, including information about continued education and scientific engagement.

**Results:** Ninety-one percent of respondents indicated that STAR U was “extremely significant” or “very significant” in influencing them to pursue a career in science, and 93% found STAR U to be effective in influencing decisions to pursue a career in aging research specifically. Forty one percent of all respondents were already accepted or enrolled in science-related advanced degree programs, with half enrolled in doctoral degree programs. Of the students who were not enrolled in graduate school, 89% of respondents indicated they had plans to enroll in advanced degrees in the future. Respondents actively disseminated their research, with 10% of STAR U scholars reporting leading or co-authoring papers intended for publication in a peer-reviewed journal (10%). In fact, review of PubMed shows that to date, 22 students (44%) have a combined total of 44 publications in peer reviewed journals. Qualitative feedback underscored the program’s impact on career exploration, as well as the impact of mentorship and the supportive environment provided by STAR U.

**Conclusions:** The STAR U program shows promise as an impactful model for advancing diversity in the scientific workforce focused on aging research by strengthening scholars’ goals for pursuing graduate education, careers in science, and research in aging in particular. Its individualized approach supports students in addressing challenges and fosters a supportive environment. STAR U serves as a catalyst for underrepresented students in STEM, showcasing the significance of tailored initiatives in promoting diversity and inclusion in aging research.

## Introduction

As the population of older adults increases dramatically, age-related illnesses such as Alzheimer’s disease and related dementias (ADRD) present a great public health burden, affecting an estimated 6.2 million.^1^ There are well documented racial/ethnic and socioeconomic disparities in incidence and prevalence of ADRD and other health outcomes in aging.^1^ Older Black and Latinx-Americans have disproportionately higher rates of ADRD, in addition to being more likely to have delayed or missed diagnoses.^2^ Innovative work led by diverse scientists, including scientists from the communities affected by these disparities, is needed to reflect the perspective and concerns of a diverse population.^3^ Yet persistent inequalities remain in the opportunities for and proportion of diverse individuals pursuing scientific careers, reflecting what Hofstra and colleagues describe as the “diversity-innovation” paradox,^5^ or the systematic devaluation of novel scientific ideas generated by scientists from minoritized backgrounds.^5^

To address the lack of scientists from diverse backgrounds, including those from groups who are underrepresented in the biomedical and behavioral sciences on a national basis, the National Institutes of Health (NIH) created specific funding opportunities to support educational activities that encourage students from such backgrounds to pursue further studies or careers in research (<u>NOT-OD-20-031: Notice of NIH’s Interest in Diversity</u>). Groups who are disproportionately under-represented in the biomedical, clinical, behavioral and social sciences include racial and ethnic groups (Blacks or African Americans, Hispanics or Latinx, American Indian or Alaska Natives, or Native Hawaiians/Pacific Islanders), individuals with disabilities, and individuals from disadvantaged backgrounds.^9^ According to the National Center for Education Statistics, while 28 percent of enrolled undergraduate students in the U.S. in 2007^7^ came from underrepresented backgrounds, only 17 percent of bachelor’s degrees in STEM fields were awarded to them.^8^ The gap is even greater with respect to graduate degrees, where students from underrepresented backgrounds earned only 13 percent of master’s degrees and 9 percent of doctoral degrees in STEM fields, compared with 18 percent of master’s degrees and 15 percent of doctoral degrees in non-STEM fields.^8^ Additionally, students from minoritized backgrounds, first-generation college students, and students from low-income backgrounds have higher attrition rates in STEM fields than their counterparts in other academic areas.^7^ While there have been recent efforts to improve the pipeline of students from backgrounds underrepresented in Science, Technology, Engineering, and Medicine (STEM) fields, more can be done to support minoritized individuals in pursuing scientific disciplines.^6^

There are several barriers to pursuing graduate degrees and scientific careers among students from underrepresented backgrounds, including limited access to mentors from similar backgrounds, economic barriers, and lack of exposure to research experiences.^10^ The development of a “research identity,” which comprises confidence, knowledge of research design, logical thinking, ability to interpret research results, and a desire to seek and succeed in research, is an important factor for academic career paths in science.^11–12^ Undergraduate summer research programs, in which students have an immersive scholarly experience as part of a cohort in an academic setting, increase retention and enrollment into doctoral programs in STEM fields and self-reported gains in research skills, writing, self-confidence, research productivity, and intellectual maturity.^13–16^ These programs provide students with intellectually stimulating learning experiences that counter the traditional classroom model while encouraging them to continue engaging in scientific research and pursue careers in STEM.^17^

Mentored, undergraduate research experiences are high impact learning practices that increase undergraduate students’ interest and preparedness for STEM careers.^18, 19^ Furthermore, aging and cognitive health training programs focused on increasing the number of scholars from minoritized backgrounds in research bring a more diverse set of experiences and perspectives to tackling the disproportionate burden of ADRD in minority communities.^20^ In 2018, the National Institute on Aging issued a funding opportunity announcement (FOA) titled “Advancing Diversity in Aging Research (ADAR)” (FOA: PAR-12-016) to help increase the diversity of the workforce committed to the aging population through undergraduate research education programs. The goals of the FOA are to increase competency in medicine, science, technology, engineering, and mathematics (MSTEM) related to aging by facilitating the transition of undergraduate students from diverse backgrounds into medicine and STEM graduate programs.

We designed the Summer of Translational Aging Research for Undergraduates (STAR U; R25 AG059557) program, which was funded in 2018 by the National Institute on Aging, in response to the above FOA. In 2021, we received additional funding from the Alzheimer’s Association to support the STAR U Program. There have been few studies that evaluate systematically the success of undergraduate training programs such as STAR U and other NIA-ADAR funded programs.^21^ Based on a recent alumni survey of the first five cohorts (2019-2023) of the STAR U program, this report describes the STAR U program and evaluates the effectiveness and success of the program in building a diverse cohort of scientists from underrepresented backgrounds.

## METHODS

### Description of the Program

STAR U provides mentored-research opportunities, complementary educational enrichment activities, and personal and professional growth experiences to undergraduate students from backgrounds historically underrepresented in the biomedical sciences. Our long-term goal is to enhance the field of aging and age-related disparities research by infusing the field with scholars from diverse and underrepresented backgrounds.

Through a structured 8-week summer research program, STAR U provides 10-14 undergraduate students per year with: 1) individual tailored research mentorships in the neuroscience of aging; 2) a range of translational learning opportunities; 3) professional networking and social experiences. Our aim is to create a learning community that is inclusive and instills a sense of belonging among students who are traditionally underrepresented in biomedical sciences. Research opportunities span areas such as patient-oriented clinical research, cognitive neuroscience and neuropsychology, basic neuroscience experimentation, and the epidemiological study of cognitive aging and its disorders. STAR U students gather formally for two-hour weekly seminar sessions that include one hour devoted to a scientific topic related to aging or neuroscience, and one hour devoted to a professional/academic development. Scientific training focuses on major themes in aging research, research methodologies, and general content areas relevant to developing scientific careers. Personal and professional development is fostered through social gatherings and journal clubs/skills-based workshops, which are led by postdoctoral fellows and cover topics such as working effectively with mentors, work-life balance, applying to graduate school, and communicating effectively in science.

Finally, the STAR U Program incorporates an empowerment-based, facilitated conversation series (STAR U Resilience Group) led by two licensed clinical psychologists who specialize in diversity and inclusion. This bi-monthly group provides support to undergraduate scholars as they navigate topics such as exploring their racial and cultural identities, navigating barriers to academic success (financial/housing/food insecurity), empowering students’ sense of belonging in the research world, and addressing mental health challenges, with the overall goal of fostering growth and resilience.

At the end of the program, students present their research at the STAR U Final Research Symposium, which takes place during the last week of the program. The symposium provides students with the opportunity to showcase their research and celebrate their growth and accomplishments during the program, in addition to learning practical skills, like scientific communication and public speaking. After the program, many students go on to present their work from STAR U at national and international scientific conferences.

### Program Eligibility

In line with the mission of the STAR U program, we encourage students from backgrounds underrepresented in the STEM disciplines to apply, based on the Notice of NIH’s Interest in Diversity.^9^ Our recruitment efforts target undergraduate diversity programs, community colleges, and minority-serving institutions. In the application, students indicate how their identity or background aligns with the goals of the program.

### Recruiting Students from Diverse and Underrepresented Backgrounds

The STAR U Program information and the online application are made publicly available through the STAR U website, which is hosted in the “Education and Training” section of <u>Columbia University’s Department of Neurology webpage.</u> The webpage provides an overview of the STAR U program, aims, highlights, program specifics, dates/deadlines, eligibility, application requirements, alumni biographies, and a list of affiliated faculty mentors and their research expertise.

To recruit STAR U applicants, the STAR U Program Coordinator reaches out to a range of undergraduate institutions including universities in the New York City metropolitan area, public and private universities across the United States, community colleges, Historically Black Colleges and Universities (HBCUs), Hispanic-serving institutions, and Tribal Colleges & Universities. Between 2019-2023, we emailed up to 560 different institutions/organizations, key individuals in various departments related to diversity and inclusion, STEM, research, and summer opportunities. In addition, our team continues to recruit and disseminate STAR U promotional information through social media channels, our website, the International Neuropsychological Society (INS) conference, Alzheimer’s Association, Society for the Advancement of Chicanos and Native Americans in Science (SACNAS), Society for Neuroscience (SfN), Annual Biomedical Research Conference for Minority Students (ABRCMS), and other non-profit organizations and societies dedicated to academic enrichment and professional development.

### Application and Selection

Students interested in applying to the STAR U program complete an online application through Qualtrics Software, which is made available on the STAR U website from November-January. All applicants are required to write a personal statement discussing their research/academic background, academic/career goals, and interest in aging research. In addition, students are asked to submit transcripts, letter(s) of recommendation, a CV/resume, and a short paragraph specifying what type of research experiences they hope to engage in over the summer. Starting in 2020, students were given the option to submit supplemental application materials in lieu of a second letter of recommendation to showcase their interests and/or skills, such as a writing sample, artwork, or video, as solicitation of multiple letters of recommendation may be a barrier for students interested in applying.^22^ Students are also asked to identify preferred mentors if accepted to STAR U and to identify their preferred type (e.g., clinical, data-driven, basic science) and topic (e.g., Alzheimer’s disease, stroke, memory) of research.

Characteristics of candidates that align with the STAR U mission include an interest in the neuroscience of aging, maturity and motivation, and an expressed interest in pursuing a career path in research. Based on frequent and in-depth selection committee (KC, SC, AMB) discussions of the submitted applications, the program coordinator contacts selected candidates for a first-round phone interview. During this call, the program coordinator shares information about STAR U and forms an initial impression about the candidate’s readiness for the program based on their ability to express and describe relevant academic and/or personal experiences to date as well as their interest in aging research. Candidates who advance to the second phase of interviews meet individually by videoconference with one of the program directors for an in-depth discussion of the candidate’s academic and scientific interests, career goals, and preferred type (e.g., clinical, data-driven, basic science) and topic (e.g., Alzheimer’s disease, stroke, memory) of research. Upon completion of second-round interviews, finalists are determined in a team meeting with the selection committee and offered acceptance to STAR U. Once students accept the offer to participate in the program, students and potential mentors are given the opportunity to meet one another via videoconference to discuss potential projects and ensure that the pairing is a good fit. Table 1 describes the application statistics from 2019-2023. An example of the STAR U application is included in the supplemental material.

**Table 1:** Application Statistics from 2019-2023.

|  | 2019 Cohort | 2020 Cohort | 2021 Cohort | 2022 Cohort | 2023 Cohort |
| --- | --- | --- | --- | --- | --- |
| Number of Applicants | 202 | 118 | 141 | 145 | 113 |
| Number of students interviewed by at least one member of the STAR U team | 50 | 26 | 26 | 39 | 24 |
| Number of final applicants selected | 10 | 10 (remote) | 13 (hybrid) | 16 (3 returning scholars) | 11 |
| Acceptance rate | 5% | 9% | 9% | 11% | 10% |

### Alumni Survey

Data were collected under Columbia University IRB protocol AAAT7823. The alumni survey was distributed between April-August 2023 to all 50 alumni of the STAR U program. The survey, administered through Qualtrics, was sent in an email describing the survey in detail, outlining that participation was voluntary, and explaining that survey results would be analyzed for presentation and publication. A study information sheet was also sent to survey respondents prior to preparation of this manuscript to ensure that alumni had the chance to opt out of the study if desired. De-identified data generated from the survey were used in the descriptive analyses reported in this paper.

The survey content included 22 questions including multiple choice, Likert-scale, and open-ended questions along with a 17-item self-reported skills assessment (See Appendix).The survey collected information on demographic data, educational and career aspirations, and a range of outcomes including graduate school admissions, published papers and presentations, impact of STAR U, and maintained interest in studying brain aging after STAR U.

## RESULTS

### STAR U Alumni Demographic Characteristics

Table 2 represents the demographic characteristics of all 50 STAR U alumni, based on our records. As indicated in Table 2, most STAR U alumni identify as women. The largest represented racial and ethnic groups were Hispanic/Latino and African American/Black, but alumni also include individuals who were Asian, American Indian or Alaska Native, individuals who identified as more than one race, and White. Most respondents majored in Neuroscience and/or Psychology, but also included Biology, Applied Math, Chemistry, Computer Science, and Human Development.

**Table 2.** Demographic Characteristics of All STAR U Alumni from 2019-2023.

|  |  |  |
| --- | --- | --- |
| **Gender** |  |  |
|  | women | 35 (70%) |
|  | men | 15 (30%) |
|  | non-binary | 0 (0%) |
| **Race/Ethnicity** |  |  |
|  | African American/Black | 16 (32%) |
|  | Hispanic/Latino (of any race) | 19 (38%) |
|  | Asian | 7 (14%) |
|  | American Indian or Alaska Native (AIAN) | 2 (4%) |
|  | More Than One Race | 3 (6%) |
|  | White | 3 (6%) |
| **Undergraduate Major** |  |  |
|  | Applied Math | 1 (2%) |
|  | Biology | 8 (18%) |
|  | Biopsychology | 1 (2%) |
|  | Chemistry | 1 (2%) |
|  | Child Development | 1 (2%) |
|  | Computer Science | 1 (2%) |
|  | Human Development | 1 (2%) |
|  | Neuroscience | 16 (36%) |
|  | Psychology | 14 (31%) |
|  | Public Health | 1 (2%) |

### Survey Responses

The 22-item survey was completed by 48 STAR U alumni, reflecting an 96% overall response rate. Table 3 outlines all survey responses. Forty-six percent of respondents reported being first-generation college students (defined as the first in their immediate family to attend undergraduate college). Almost half of the survey respondents (48%) reported working in a position or pursuing studies related to brain aging.

**Table 3.**
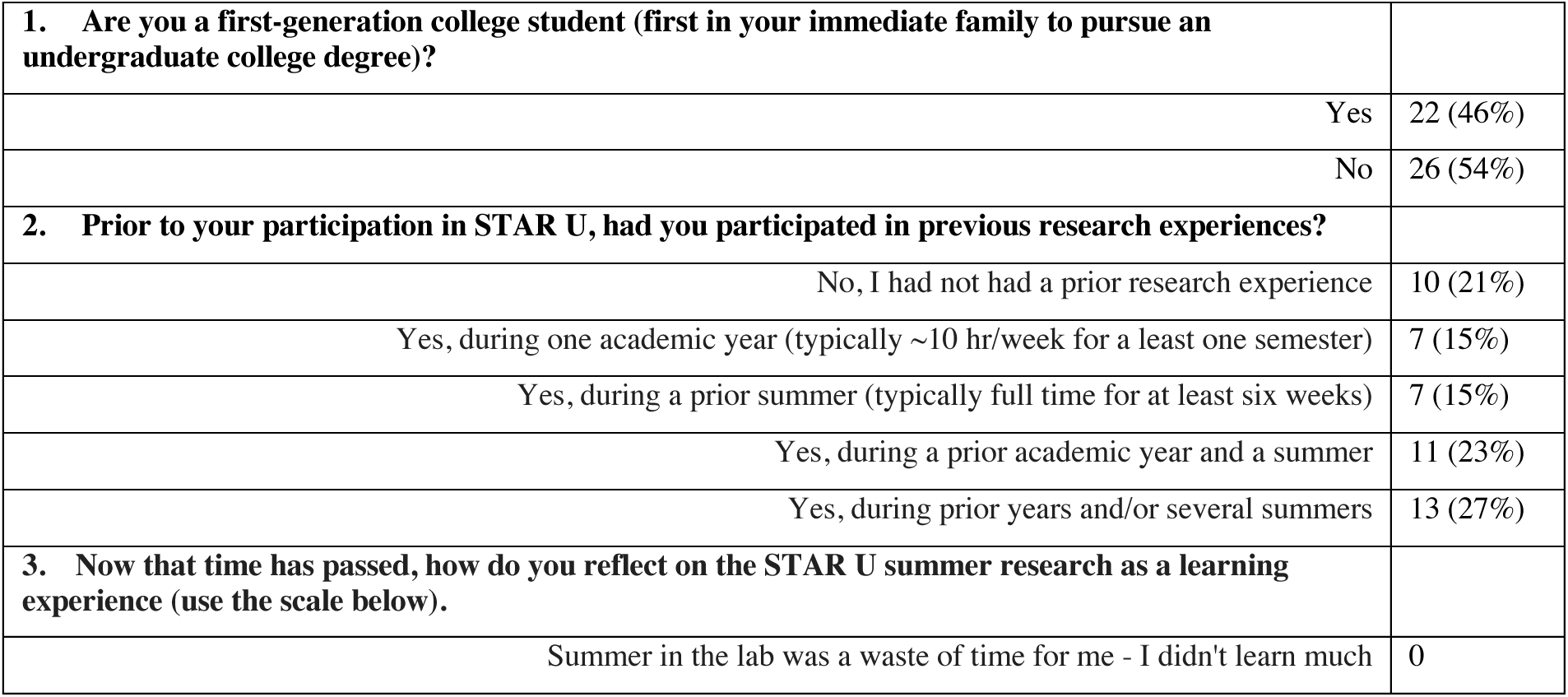
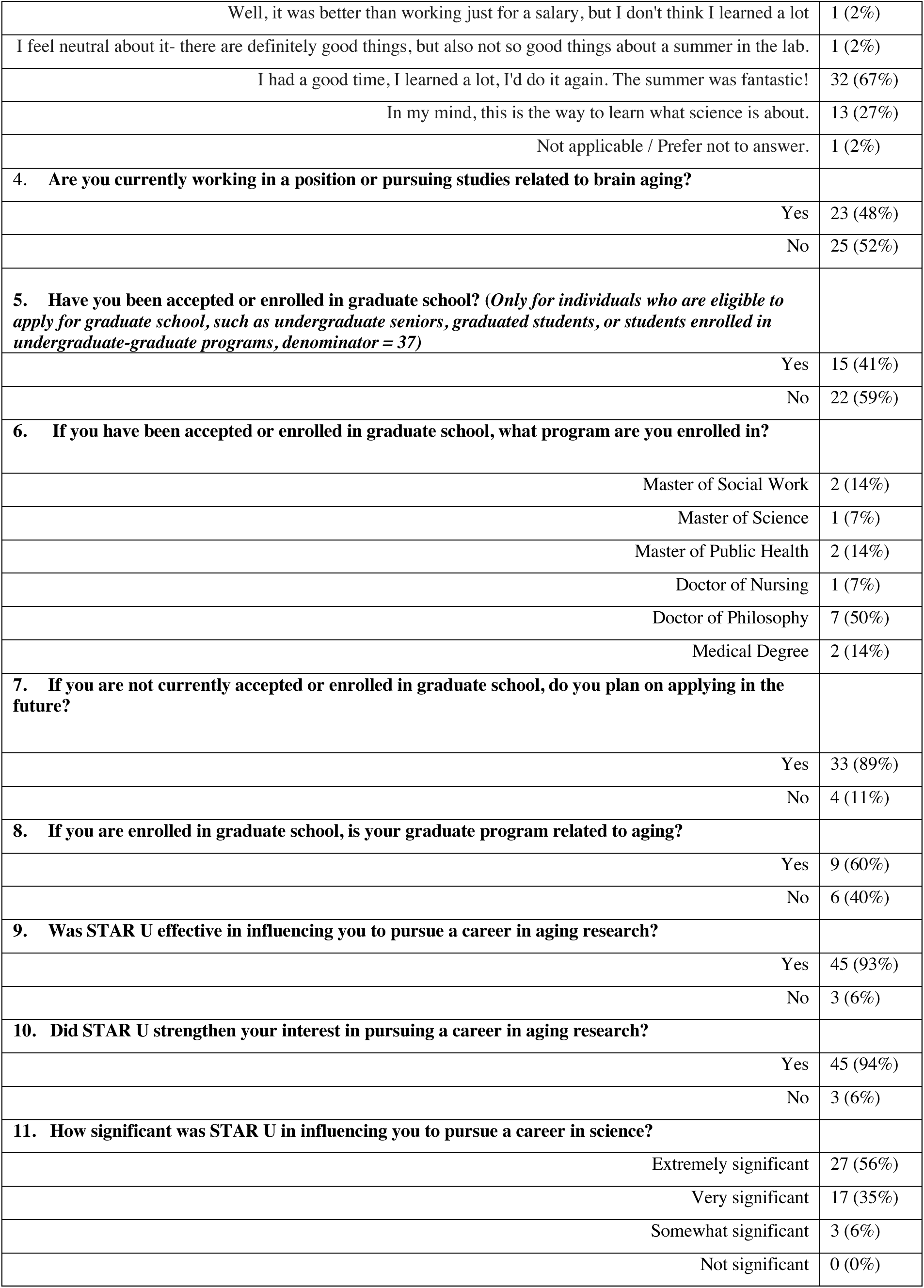
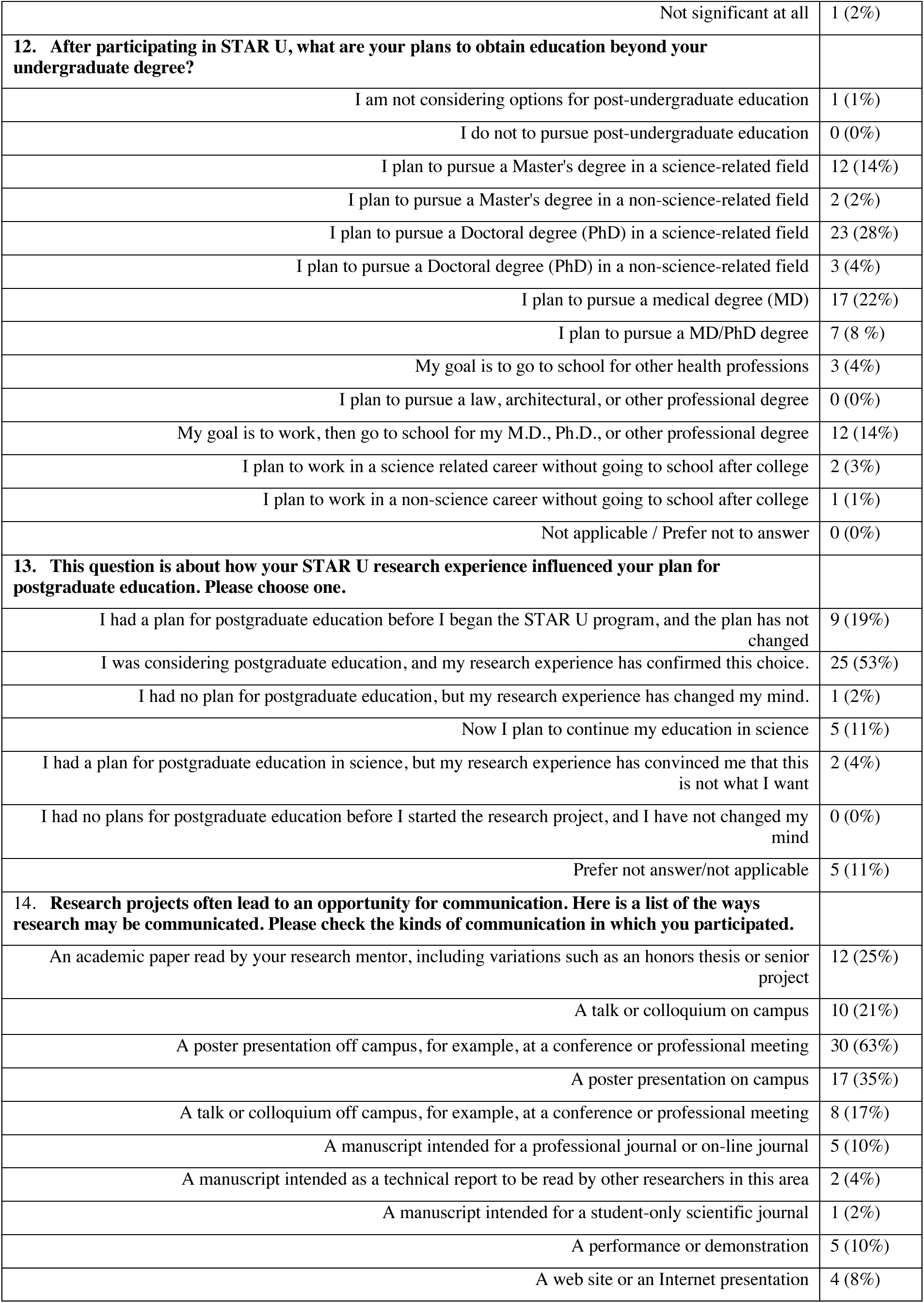
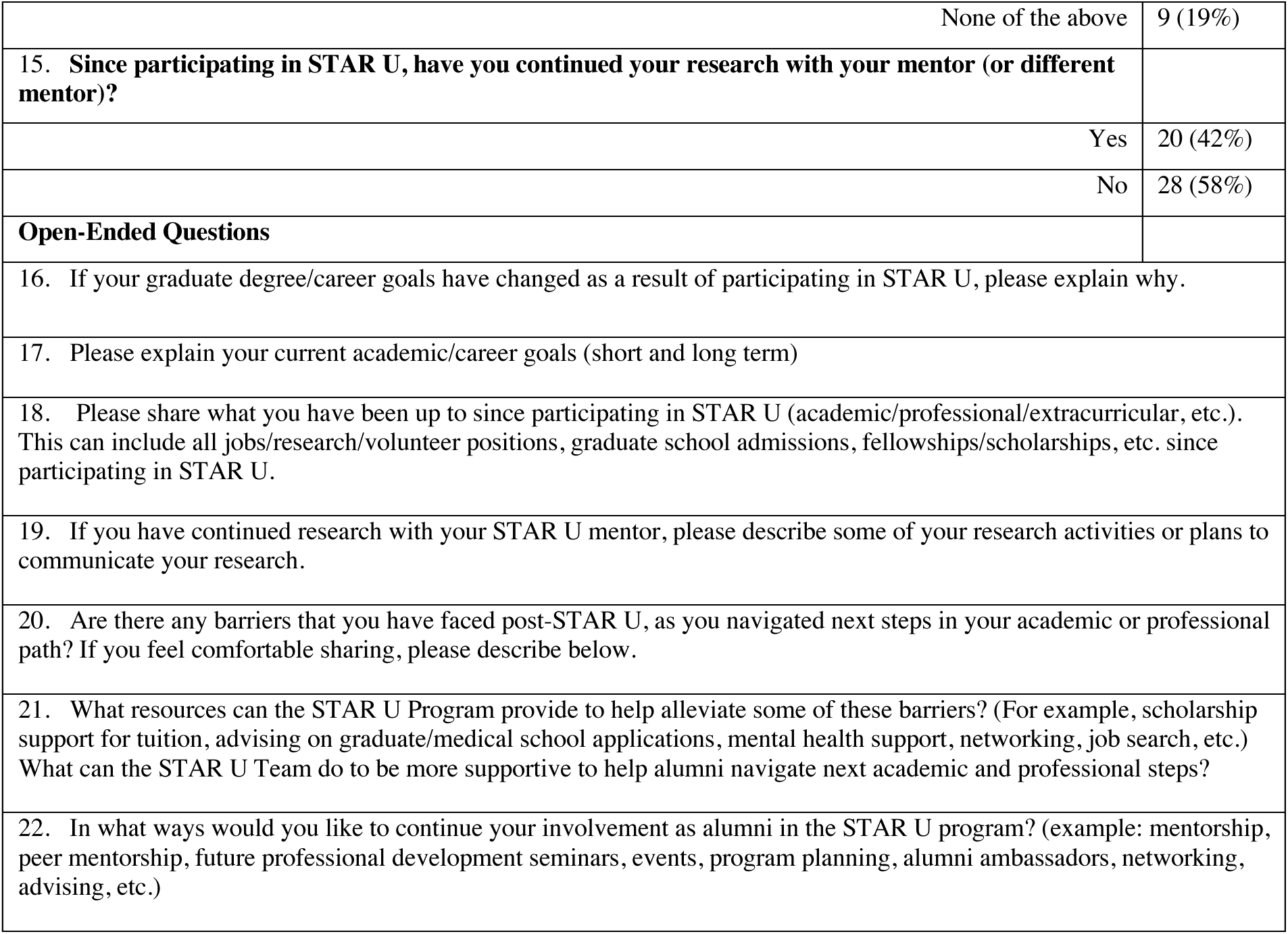
Survey Responses of STAR U Alumni from 2019-2023.

|  |  |  |
| --- | --- | --- |
| **1. Are you a first-generation college student (first in your immediate family to pursue an undergraduate college degree)?** |  |  |
|  | Yes | 22 (46%) |
|  | No | 26 (54%) |
| **2. Prior to your participation in STAR U, had you participated in previous research experiences?** |  |  |
|  | No, I had not had a prior research experience | 10 (21%) |
|  | Yes, during one academic year (typically ~10 hr/week for a least one semester) | 7 (15%) |
|  | Yes, during a prior summer (typically full time for at least six weeks) | 7 (15%) |
|  | Yes, during a prior academic year and a summer | 11 (23%) |
|  | Yes, during prior years and/or several summers | 13 (27%) |
| **3. Now that time has passed, how do you reflect on the STAR U summer research as a learning experience (use the scale below).** |  |  |
|  | Summer in the lab was a waste of time for me - I didn't learn much | 0 |

|  |  |
| --- | --- |
| Well, it was better than working just for a salary, but I don't think I learned a lot | 1 (2%) |
| I feel neutral about it- there are definitely good things, but also not so good things about a summer in the lab. | 1 (2%) |
| I had a good time, I learned a lot, I'd do it again. The summer was fantastic! | 32 (67%) |
| In my mind, this is the way to learn what science is about. | 13 (27%) |
| Not applicable / Prefer not to answer. | 1 (2%) |
| **4. Are you currently working in a position or pursuing studies related to brain aging?** |  |
| Yes | 23 (48%) |
| No | 25 (52%) |
| **5. Have you been accepted or enrolled in graduate school? (Only for individuals who are eligible to apply for graduate school, such as undergraduate seniors, graduated students, or students enrolled in undergraduate-graduate programs, denominator = 37)** |  |
| Yes | 15 (41%) |
| No | 22 (59%) |
| **6. If you have been accepted or enrolled in graduate school, what program are you enrolled in?** |  |
| Master of Social Work | 2 (14%) |
| Master of Science | 1 (7%) |
| Master of Public Health | 2 (14%) |
| Doctor of Nursing | 1 (7%) |
| Doctor of Philosophy | 7 (50%) |
| Medical Degree | 2 (14%) |
| **7. If you are not currently accepted or enrolled in graduate school, do you plan on applying in the future?** |  |
| Yes | 33 (89%) |
| No | 4 (11%) |
| **8. If you are enrolled in graduate school, is your graduate program related to aging?** |  |
| Yes | 9 (60%) |
| No | 6 (40%) |
| **9. Was STAR U effective in influencing you to pursue a career in aging research?** |  |
| Yes | 45 (93%) |
| No | 3 (6%) |
| **10. Did STAR U strengthen your interest in pursuing a career in aging research?** |  |
| Yes | 45 (94%) |
| No | 3 (6%) |
| **11. How significant was STAR U in influencing you to pursue a career in science?** |  |
| Extremely significant | 27 (56%) |
| Very significant | 17 (35%) |
| Somewhat significant | 3 (6%) |
| Not significant | 0 (0%) |
| Not significant at all | 1 (2%) |
| **12. After participating in STAR U, what are your plans to obtain education beyond your undergraduate degree?** |  |
| I am not considering options for post-undergraduate education | 1 (1%) |
| I do not to pursue post-undergraduate education | 0 (0%) |
| I plan to pursue a Master's degree in a science-related field | 12 (14%) |
| I plan to pursue a Master's degree in a non-science-related field | 2 (2%) |
| I plan to pursue a Doctoral degree (PhD) in a science-related field | 23 (28%) |
| I plan to pursue a Doctoral degree (PhD) in a non-science-related field | 3 (4%) |
| I plan to pursue a medical degree (MD) | 17 (22%) |
| I plan to pursue a MD/PhD degree | 7 (8 %) |
| My goal is to go to school for other health professions | 3 (4%) |
| I plan to pursue a law, architectural, or other professional degree | 0 (0%) |
| My goal is to work, then go to school for my M.D., Ph.D., or other professional degree | 12 (14%) |
| I plan to work in a science related career without going to school after college | 2 (3%) |
| I plan to work in a non-science career without going to school after college | 1 (1%) |
| Not applicable / Prefer not to answer | 0 (0%) |
| **13. This question is about how your STAR U research experience influenced your plan for postgraduate education. Please choose one.** |  |
| I had a plan for postgraduate education before I began the STAR U program, and the plan has not changed | 9 (19%) |
| I was considering postgraduate education, and my research experience has confirmed this choice. | 25 (53%) |
| I had no plan for postgraduate education, but my research experience has changed my mind. | 1 (2%) |
| Now I plan to continue my education in science | 5 (11%) |
| I had a plan for postgraduate education in science, but my research experience has convinced me that this is not what I want | 2 (4%) |
| I had no plans for postgraduate education before I started the research project, and I have not changed my mind | 0 (0%) |
| Prefer not answer/not applicable | 5 (11%) |
| **14. Research projects often lead to an opportunity for communication. Here is a list of the ways research may be communicated. Please check the kinds of communication in which you participated.** |  |
| An academic paper read by your research mentor, including variations such as an honors thesis or senior project | 12 (25%) |
| A talk or colloquium on campus | 10 (21%) |
| A poster presentation off campus, for example, at a conference or professional meeting | 30 (63%) |
| A poster presentation on campus | 17 (35%) |
| A talk or colloquium off campus, for example, at a conference or professional meeting | 8 (17%) |
| A manuscript intended for a professional journal or on-line journal | 5 (10%) |
| A manuscript intended as a technical report to be read by other researchers in this area | 2 (4%) |
| A manuscript intended for a student-only scientific journal | 1 (2%) |
| A performance or demonstration | 5 (10%) |
| A web site or an Internet presentation | 4 (8%) |
| None of the above | 9 (19%) |
| **15. Since participating in STAR U, have you continued your research with your mentor (or different mentor)?** |  |
| Yes | 20 (42%) |
| No | 28 (58%) |
| **Open-Ended Questions** |  |
| 16. If your graduate degree/career goals have changed as a result of participating in STAR U, please explain why. |  |
| 17. Please explain your current academic/career goals (short and long term) |  |
| 18. Please share what you have been up to since participating in STAR U (academic/professional/extracurricular, etc.). This can include all jobs/research/volunteer positions, graduate school admissions, fellowships/scholarships, etc. since participating in STAR U. |  |
| 19. If you have continued research with your STAR U mentor, please describe some of your research activities or plans to communicate your research. |  |
| 20. Are there any barriers that you have faced post-STAR U, as you navigated next steps in your academic or professional path? If you feel comfortable sharing, please describe below. |  |
| 21. What resources can the STAR U Program provide to help alleviate some of these barriers? (For example, scholarship support for tuition, advising on graduate/medical school applications, mental health support, networking, job search, etc.) What can the STAR U Team do to be more supportive to help alumni navigate next academic and professional steps? |  |
| 22. In what ways would you like to continue your involvement as alumni in the STAR U program? (example: mentorship, peer mentorship, future professional development seminars, events, program planning, alumni ambassadors, networking, advising, etc.) |  |

### Graduate School Outcomes

Forty-one percent of all respondents eligible to apply for graduate school reported acceptance or enrollment in graduate school at the time of the survey. For those currently accepted or enrolled in graduate school, diverse degree programs were represented, with half of the respondents pursuing doctoral degrees. Other respondents indicated acceptance or enrollment in programs to obtain Masters of Social Work, Masters of Science, Masters of Public Health, and Medical Degrees, as summarized in Table 3.

Among respondents not currently accepted or enrolled in graduate school, the majority expressed intentions to apply for further education in the future (89%). Conversely, a smaller portion of respondents indicated they did not plan to pursue graduate education, citing reasons such as a desire to engage in full-time work to support their families financially or to prevent burnout.

### Influence of STAR U on Career and Educational Goals

With regard to the influence of STAR U on career and educational goals, most respondents (93%) reported that STAR U was effective in influencing their decision to pursue careers in aging research. When asked how significant STAR U was in influencing respondents’ pursuit of careers in science more broadly, most respondents indicated that STAR U was “extremely significant” (56%) or “very significant” (35%).

### Impact on Postgraduate Educational Plans

Analysis of the impact of STAR U on postgraduate education plans revealed that half of respondents had already considered postgraduate education before joining the program. Their experience in STAR U reaffirmed this choice, while a much smaller percentage, who initially had no plans for graduate school, began considering postgraduate education after completing the program (4%). Very few respondents who initially planned for postgraduate education changed their minds due to their research experience (4%).

### Research Communication and Continuation

Survey respondents engaged in various forms of dissemination of their STAR U research beyond participating in the program. STAR U scholars participated in various forms of research dissemination, including poster presentations or talks at off-campus and on-campus conferences, including the Advancing Diversity for Aging Research Summit, the Gerontological Society of America annual meeting, the Advancing Biomedical Research Conference for Minority Students, the Alzheimer’s Association International Conference, and the International Neuropsychological Society annual meeting. See Table 3. Moreover, some STAR U scholars reported leading or co-authoring papers intended for publication in a peer-reviewed journal (10%). In fact, review of PubMed shows that to date, 22 students (44%) have a combined total of 44 publications in peer reviewed journals. Finally, respondents reported ongoing research with their mentors (or different mentors) after4 participating in STAR U (Table 3).

### Skills-Based Assessment

The skills-based assessment (Table 4) indicated that most respondents reported a “very large gain,” “large gain,” or “moderate gain” in every skills/benefits category, demonstrated by the darker shades of green in the table. The percentage of students indicating a “small gain” or “no gain” was minimal, to none. Skills with the largest gain included “becoming part of a learning community,” (71%), “readiness for more demanding research” (54%), and “understanding the research process” (54%).

**Table 4.** Self-Reported Skills, STAR U Alumni from 2019-2023.

| After participating in STAR U, please consider some of the possible benefits you may have gained from your research experience. |  |  |  |  |  |  |
| --- | --- | --- | --- | --- | --- | --- |
|  | No gain or very small gain | Small gain | Moderate gain | Large gain | Very large gain | N/A or prefer not to answer |
| Learning to work independently | 2% | 2% | 27% | 29% | 37% | 2% |
| Readiness for more demanding research | 4% | 4% | 10% | 27% | 54% | 0 |
| Ability to integrate theory and practice | 2% | 8% | 27% | 21% | 40% | 2% |
| Skill in scientific writing | 0 | 12% | 23% | 27% | 33% | 4% |
| Skill in giving an effective oral presentation | 8% | 0 | 17% | 31% | 42% | 2% |
| Self confidence in research | 4% | 6% | 12% | 29% | 49% | 0 |
| Learning ethical conduct in your field | 2% | 8% | 14% | 33% | 37% | 4% |
| Ability to read and understand primary literature | 2% | 2% | 23% | 27% | 42% | 4% |
| Ability to analyze data and other information | 2% | 4% | 21% | 37% | 31% | 4% |
| Learning laboratory techniques | 0 | 10% | 27% | 12% | 27% | 17% |
| Skill in interpreting results | 2% | 2% | 25% | 33% | 33% | 4% |
| Understanding how knowledge is constructed | 2% | 4% | 12% | 35% | 44% | 2% |
| Tolerance for obstacles faced in research | 2% | 4% | 10% | 33% | 48% | 2% |
| Understanding the research process | 0 | 6% | 12% | 27% | 54% | 0 |
| Confidence in your potential to be a scientist | 2% | 0 | 19% | 29% | 50% | 0 |
| Clarification of a career path | 2% | 0 | 10% | 33% | 52% | 2% |
| Becoming part of a learning community | 0 | 4% | 4% | 21% | 71% | 0 |

\*Darker shades of green indicate higher concentration of students in that scale category and lighter shades of green/white indicate lower concentration of responses in that category.

### Qualitative Results

The qualitative feedback from STAR U respondents illuminated several key themes, including opportunities for career exploration and academic growth, mentorship, and safe spaces for growth. These themes were determined by the first author (KC), who reviewed the qualitative transcripts several times and made note of key themes that emerged.

### Career Exploration and Academic Growth

Respondents underscored their growth, with one noting, “I grew as a researcher and learned more about what it takes to go into grad school.” Some respondents expressed newfound interests. As one participant stated, “I feel more prepared for my future in the field and gained a real interest in the field of aging research.” Shadowing experiences were highlighted in the responses of several scholars, “This program allowed me to have a wonderful experience to complete work on the research side of things but also get shadowing experiences.” Exposure to both clinical and research careers through shadowing was frequently mentioned, as articulated by one participant: “I got to see how solely from a patient’s presentation, the stroke team could infer what type of neurological condition the patient was experiencing.” The program also contributed to skills development, with one participant expressing, “Working with data for my project helped me gain coding literacy.” Respondents indicated their growth in various research-related skills, as demonstrated in Table 4.

### Mentorship

STAR U Scholars highlighted the importance of mentorship during their summer experience. As one participated stated, “Both of my mentors were very supportive and helpful in guiding me with my research project. There were many times where I did feel lost about my research, and they helped clarify any doubts I had. My mentors encouraged me to ask questions, were very understanding, and helped any way they could.” Qualitative responses also highlighted the importance of mentors’ availability to teach research methodology, discuss projects, and keep respondents on track throughout the program. Moreover, students valued the fact that mentors emphasized the program as a learning experience, fostered a supportive and approachable environment, and encouraged respondents through various stages of their projects.

As one respondent stated: “My faculty mentor was amazing! They were extremely helpful and so involved in my project every step of the way! I appreciated how much time they dedicated to the program and how accessible they were to reach when I was having trouble grasping certain concepts. I loved how much they emphasized that this was a learning experience and that I should not feel in any way like I needed to have everything down right away which made me feel very supported and comfortable in this environment.”

Beyond support in research methodology, respondents appreciated their mentors’ approachability, patience in explaining concepts, willingness to engage in broader discussions, flexibility and understanding. Mentors’ willingness to accommodate discussions outside regular work hours and their understanding attitude towards respondents’ mental health were also underscored in survey responses.

### Safe Spaces for Growth

Survey participants were asked to share their opinions of the STARU Resilience Group. Post-program survey responses from the resilience group between 2021-2023 can be found in the appendix. One participant noted, “Having the opportunity to widen our understanding of how microaggressions, imposter syndrome, and burnout can affect different people is so important.” Qualitative responses characterized the group as non-judgmental, fostering open and honest communication. Respondents expressed that, “at the end of each session, they always let us know we could reach out to them for help.”

## DISCUSSION

NIH’s mission is to advance scientific knowledge and improve public health, recognizing that achieving these goals requires a workforce that reflects the diversity of the population it serves.^24^ STAR U is aligned with this mission, striving to nurture and train aspiring scientists from underrepresented backgrounds in brain aging research. STAR U has succeeded in engaging students from a range of underrepresented backgrounds in STEM. Most scholars identified as Hispanic/Latino, African American/Black, and Asian, with a large proportion of students identifying as first-generation college attendees. This diversity mirrors that which is needed to address the persistent lack of diversity in STEM fields, where opportunities for mentorship and research exposure are often limited.^25^

STAR U consistently attracts motivated and ambitious students who proactively seek admission to the program. Despite the absence of a comparison group to assess the counterfactual scenario – i.e., how students would have fared without program participation – we bridge this gap by conducting alumni surveys. These surveys delve into the extent to which students see STAR U as influential in their educational and career plans, providing valuable insights to deepen our understanding of the program’s impact.

Formal evaluations from STAR U alumni spanning the first five years of the program provide promising indicators of the positive impact that STAR U has had in supporting the academic, career, and personal pursuits of scholars. First, the impact of STAR U on postgraduate education plans is notable. The majority of the respondents confirmed that their research experience in STAR U either reinforced their pre-existing plan to pursue postgraduate education or confirmed previous considerations about graduate education, suggesting that STAR U effectively encouraged respondents to pursue their educational ambitions. Furthermore, a small percentage of students who initially planned to attend graduate school for science-related degrees decided to change their paths. Although these changes may not have aligned with the original goals of the program, they highlight the value of research experiences in helping students refine their career goals.

One of the most notable findings from the alumni survey was the influence of the STAR U program on respondents’ career aspirations. The vast majority of respondents indicated that STAR U was extremely or very significant in influencing them to pursue a career in science more broadly, and effective in influencing their decision to pursue careers in aging research specifically. With regard to specific degree programs, a large proportion of students expressed an intention to pursue doctoral degrees in science-related fields and a substantial number of students aspired to attend medical school or obtain master’s degrees, reflecting the interdisciplinary nature of the program. Among STAR U alumni who are eligible to apply to graduate school including undergraduate seniors, graduated students, or students enrolled in undergraduate-graduate programs (n = 37), 41% were accepted to or enrolled in graduate programs. This rate is more than three times the national average among individuals from underrepresented backgrounds, or 13% earning master’s degrees, and only 9% of doctoral degrees.^8^ Our survey revealed that respondents of STAR U actively engaged in research communication through poster presentations, academic papers, and talks, showcasing their research outcomes at national and international conferences and on-campus events.

The skills-based assessment revealed substantial gains experienced by respondents across various skills and benefits categories. The prevalence of “very large gain,” “large gain,” or “moderate gain” underscores the robustness of the STAR U curriculum, and its effectiveness in building skill development among participants. In particular, key skills/benefits such as being part of a learning community, readiness for more demanding research, and a deeper understanding of the research process, emerged as areas where respondents experienced the most significant growth. These findings point to STAR U’s potential in nurturing a supportive learning environment while enhancing participants’ capabilities to engage in rigorous research activities. Our results underscore the importance of targeted curriculum in promoting skill acquisition and preparing individuals for academic and professional pursuits.

Furthermore, qualitative results demonstrated that academic/career growth opportunities, mentorship, and a supportive environment were pivotal aspects of the STAR U program, shaping scholars’ research experiences throughout the summer. Through immersive shadowing opportunities, seminar didactics, and STAR U Resilience Group sessions, students indicated gaining valuable insights into both research and clinical aspects of the field, fostering a deeper appreciation for aging-related research and its potential impact. Respondents shared the invaluable support and guidance received from their mentors, who played a central role in helping students overcome their doubts and offering support throughout the summer. The continuation of research with mentors beyond the summer program also underscores the strong mentorship component of STAR U, as students develop long-lasting connections with their mentors, relationships which are crucial for sustained engagement in research.

Finally, dedicated spaces in which to foster a supportive community, such as the STAR U Resilience Group, emerged as a vital resource for scholars, providing a safe and inclusive space for discussions on topics such as microaggressions, imposter syndrome, and burnout. Facilitators were commended for their commitment to fostering open and honest dialogue, and creating a sense of belonging and support among participants. The provision of ongoing assistance and encouragement further solidified the group’s importance in nurturing participants’ emotional resilience and well-being.

Survey data demonstrate that mentored summer research experiences such as STAR U, can be transformative in guiding young, diverse minds toward scientific careers. An area for improvement includes creating additional engagement opportunities with alumni and the STAR U community in the form of alumni seminars, check-ins, and peer mentoring. This engagement would aim to support alumni beyond their participation in the STAR U program.

## Conclusion

In light of the aging U.S. population and the increasing demand for research in aging-related diseases, it is imperative to nurture diverse and talented scientists.^26,27^ The Summer of Translational Aging Research for Undergraduates (STAR U) program has promise as an effective training model for advancing diversity within the realm of ADRD research. Its reported success in supporting and shaping scholars’ career aspirations, educational objectives, and research engagement underscores the pivotal role such programs play in addressing disparities within STEM fields. STAR U focuses on the individual needs of scholars, fostering an environment where students can learn in a nurturing, supportive environment that addresses some of the challenges students may face, such as being a first-generation college student, low-income, or being underrepresented in their fields. The program not only serves as a driving force for underrepresented students in STEM, but also exemplifies how tailored summer research initiatives can be crucial in promoting diversity, equity, and inclusion, especially within the intricate field of aging research.^28^

Regarding limitations of the current study, while we had a high overall response rate, the response data may bias towards more positive feedback compared to non-response data.

Furthermore, as many STAR U alumni are still undergraduates in college, it will take more time to conduct a more comprehensive understanding of the longer-term impact of STAR U. The mid-term effects of STAR U are evident in the positive trajectories of its alumni, signaling a promising pathway toward achieving greater diversity and fostering innovation within aging research and beyond. As we navigate the future landscape of STEM, we should keep in mind the importance of holistic, mentored summer research programs that can create meaningful and lasting impacts on the scientific community.

## Data Availability

All data produced in the present study are available upon reasonable request to the authors

## Acknowledgements and Disclosures

Acknowledgements

The authors gratefully acknowledge the support of the National Institute on Aging Advancing Diversity in Aging Research through Undergraduate Education (ADAR) program for funding the STAR U program. Additionally, we extend our appreciation to all mentors and facilitators whose dedication to student mentoring has been invaluable in shaping the STAR U community.

## Funding/Support

National Institute of Aging R25 Grant, STAR U, Grant # AG059557

Other Disclosures

Not applicable

Ethical Approval

Columbia University IRB protocol AAAT7823

Disclaimers

Not applicable

Previous Presentations

Not applicable

## Resilience Group Questions 2021 and 2023 (Average)

On a scale of 1-5, with 1 being “Strongly Disagree” and 5 being “Strongly Agree,” how much do you agree with this statement: I am aware of the negative impact of racial battle fatigue on your neurological, mental, and physical well-being.

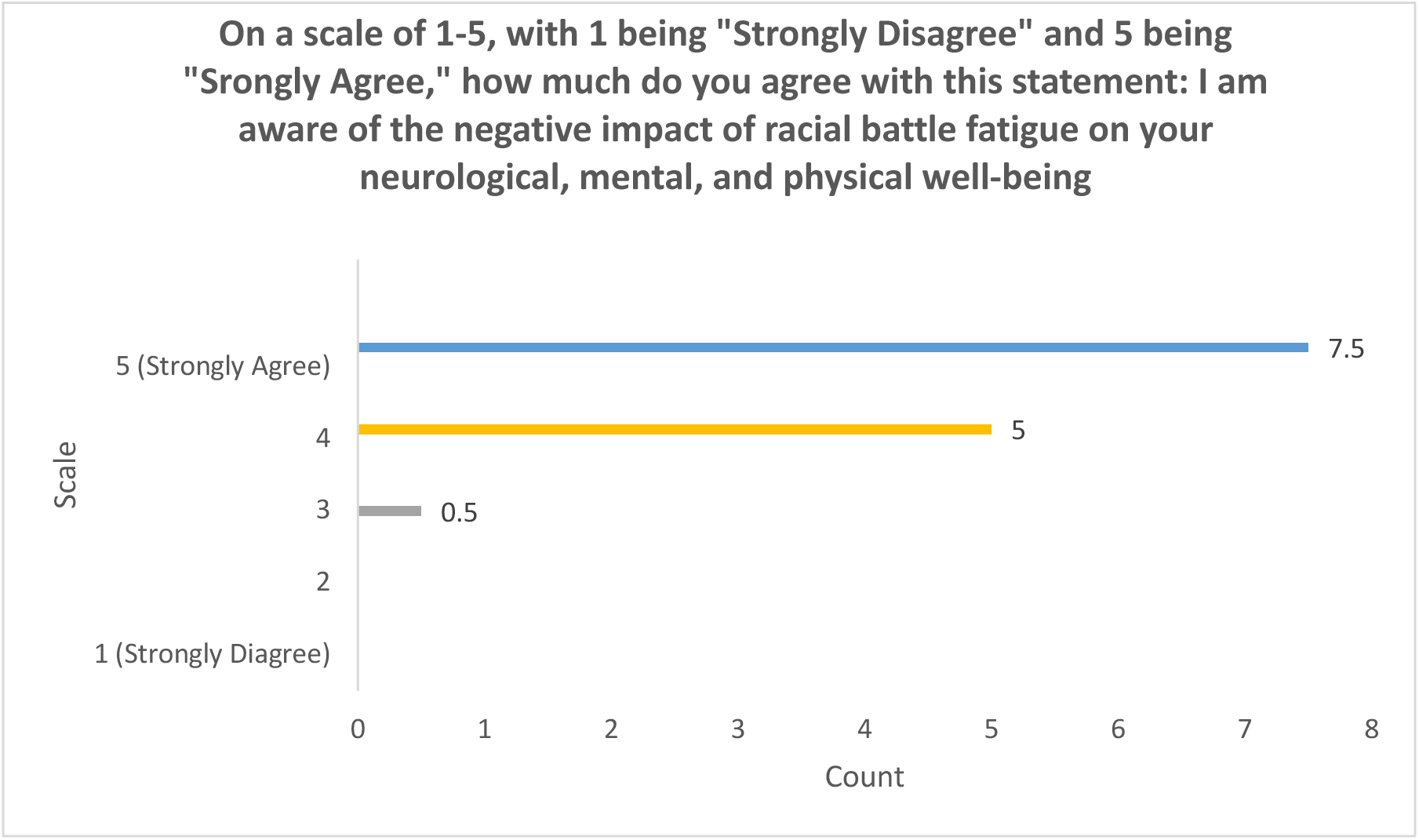

On a scale of 1-5, with 1 being “Strongly Disagree” and 5 being “Strongly Agree,” how much do you agree with this statement: I am aware of how to identify my values and align them with my behavior.

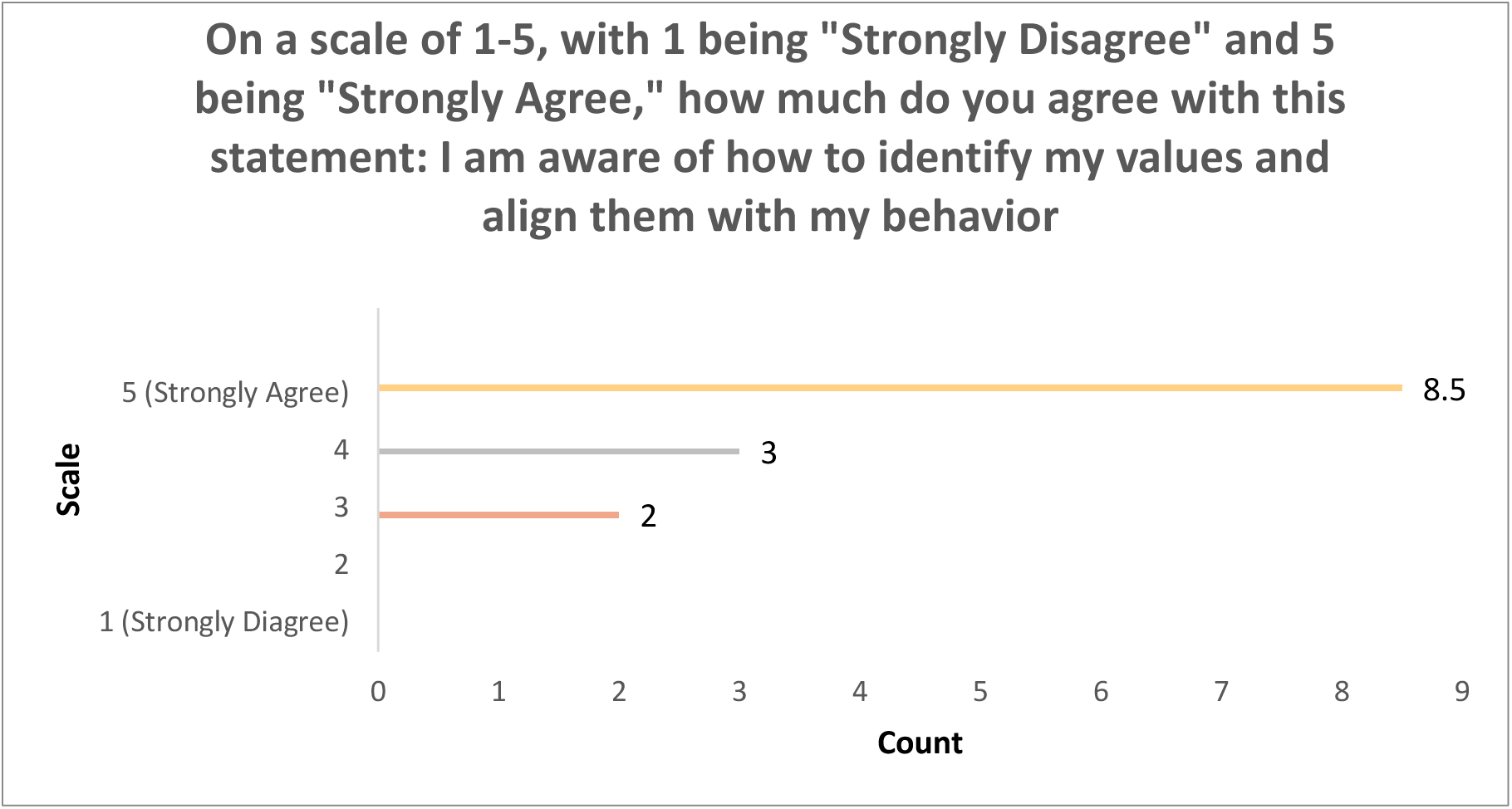

As a result of the STAR U Resilience group, I feel more connected to other students who understand me and my experience.

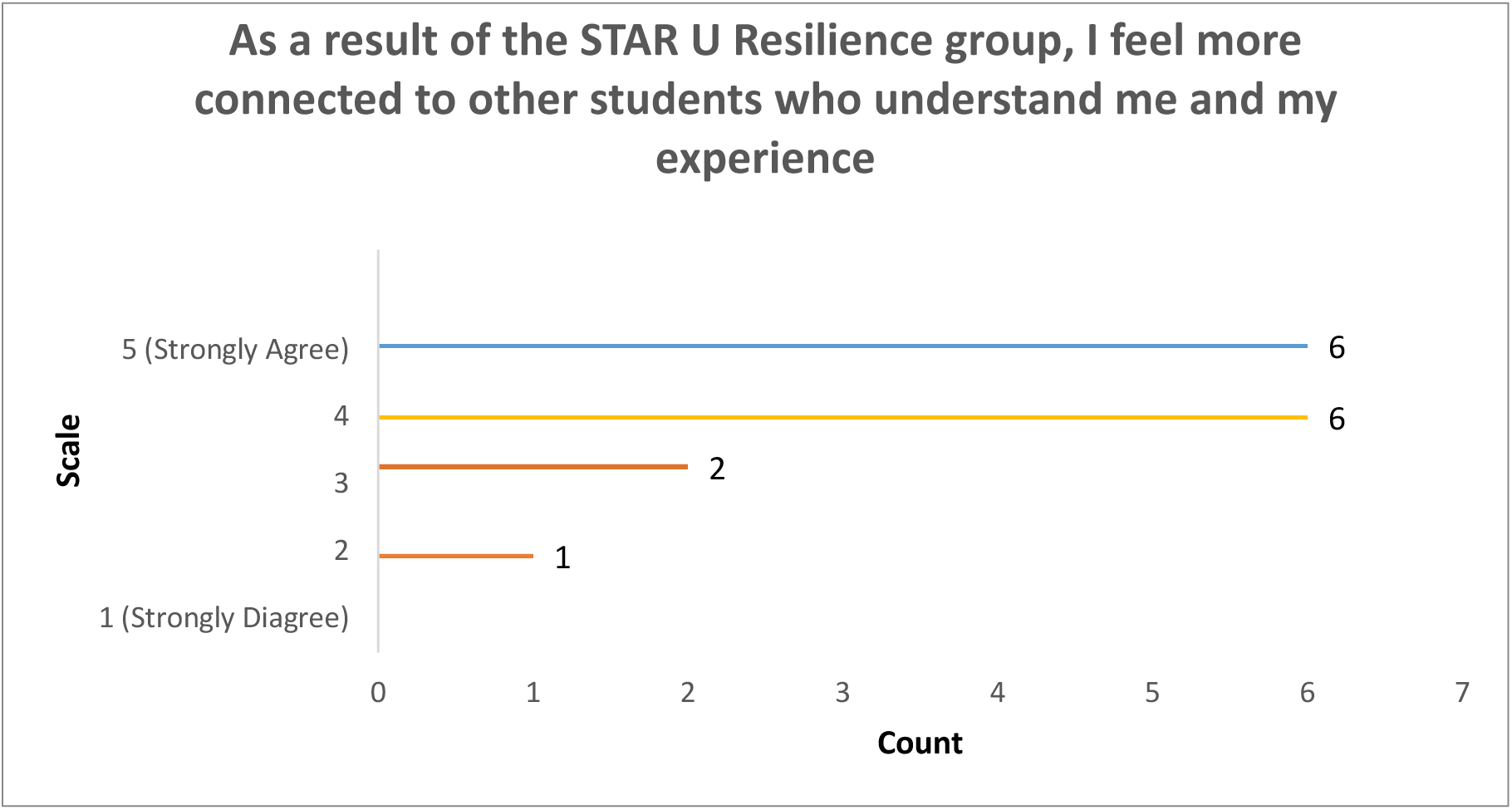

This group provided me with the type of mental health support needed to continue working in the research field.

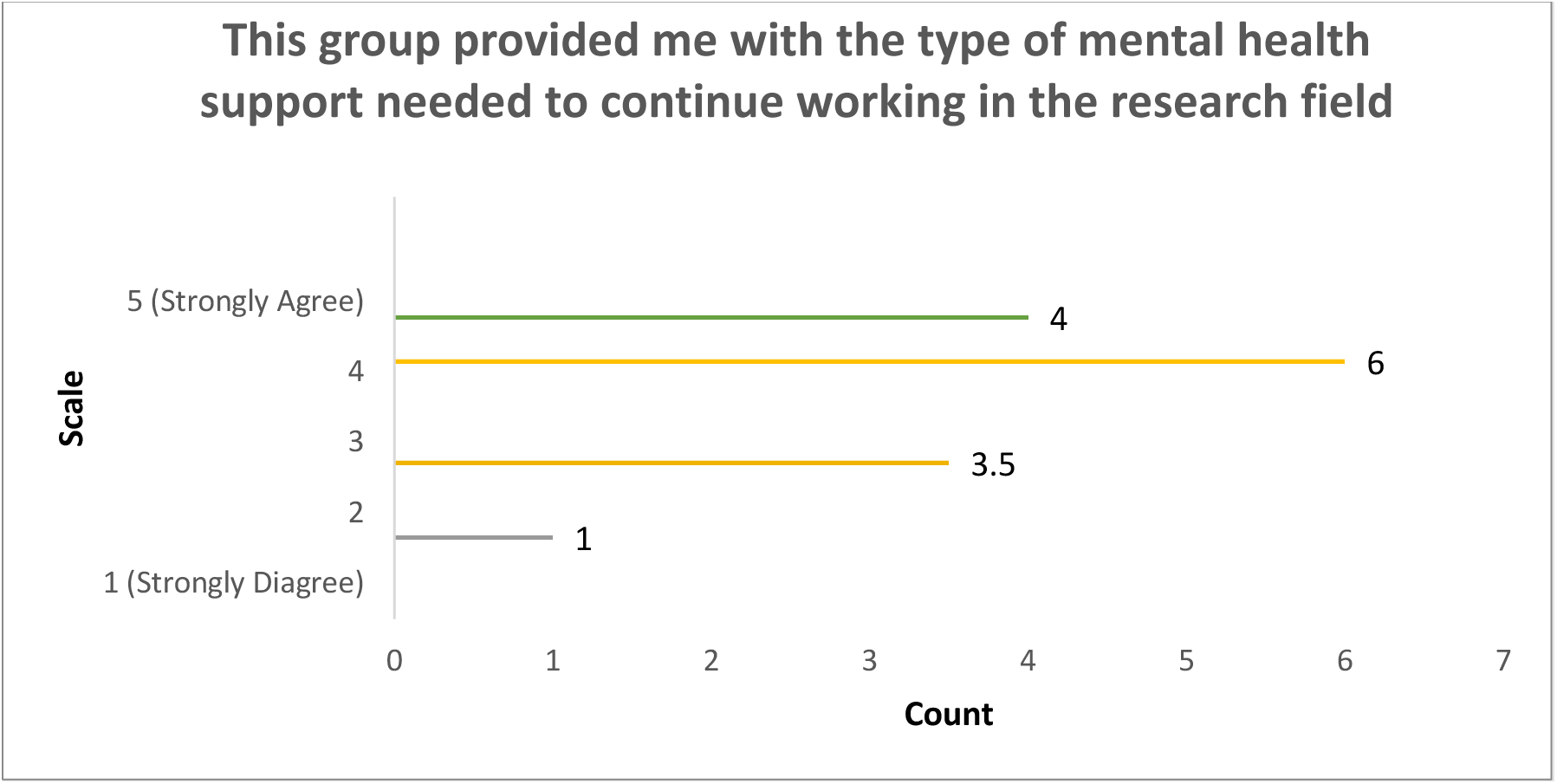

How important was it for the STAR U Resilience Group to be led by people of color mental health professionals?

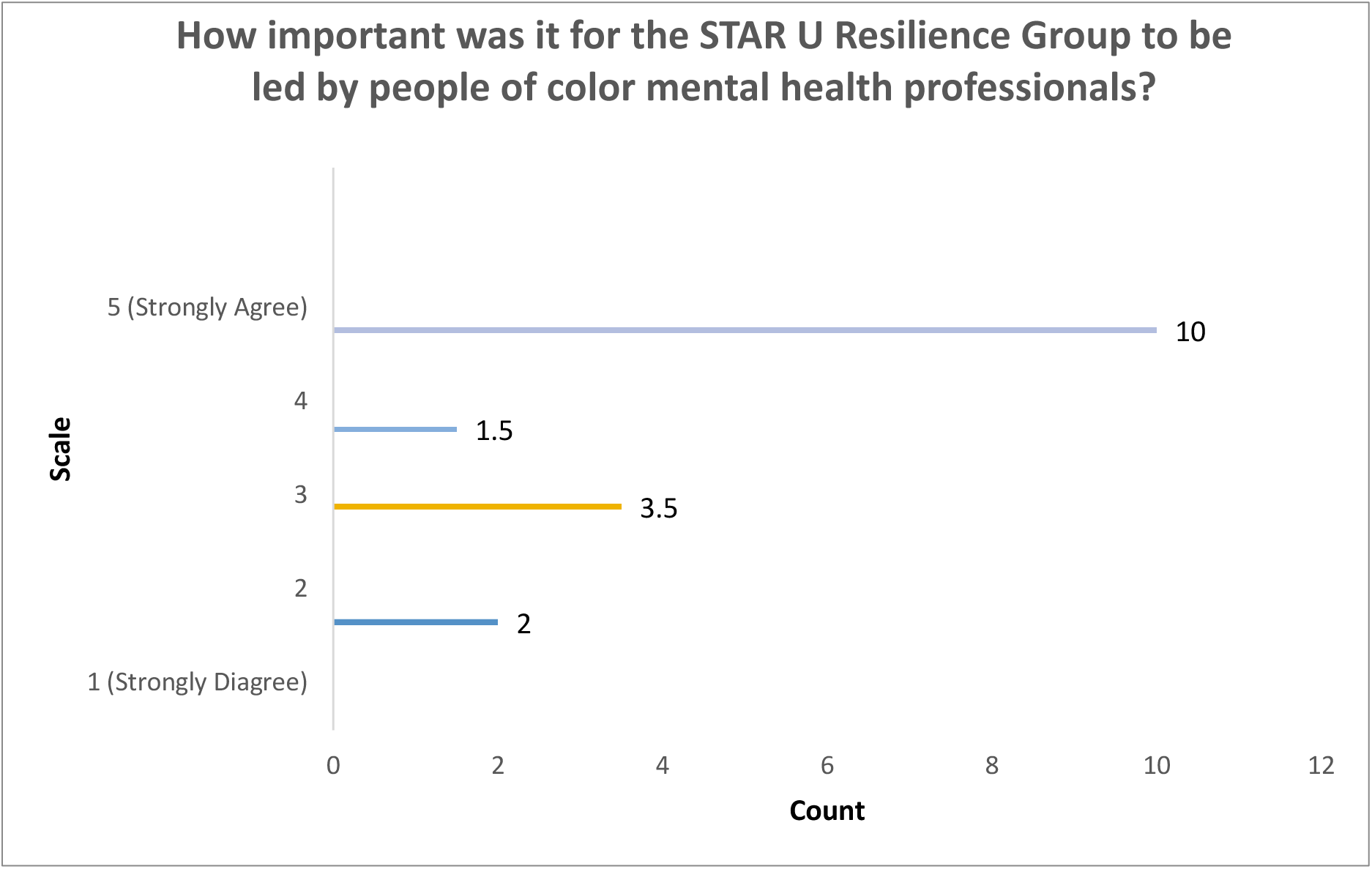

As a result of this group, I felt seen, valued, validated, and empowered.

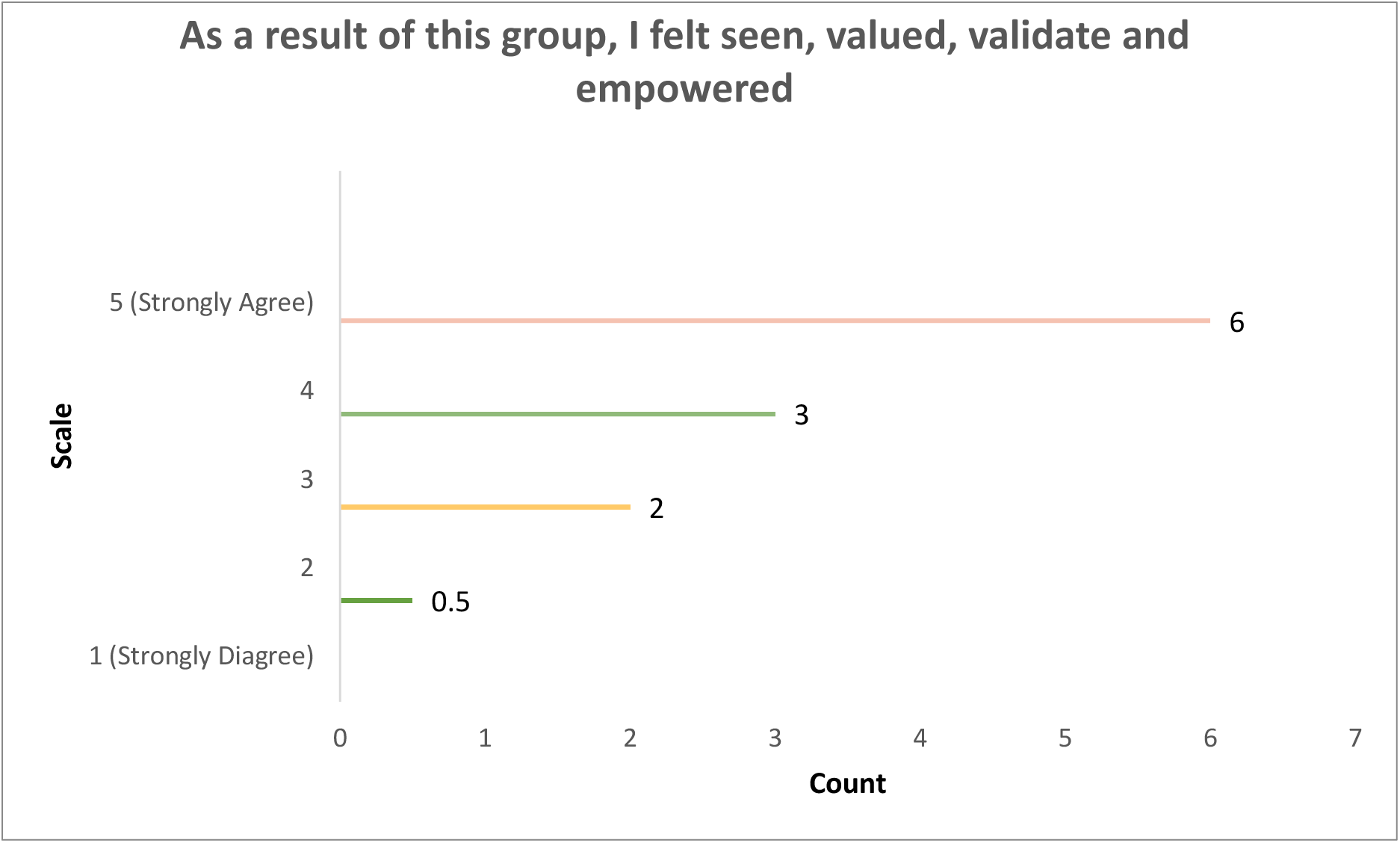

